# Estimation of COVID-2019 burden and potential for international dissemination of infection from Iran

**DOI:** 10.1101/2020.02.24.20027375

**Authors:** Ashleigh R. Tuite, Isaac I. Bogoch, Ryan Sherbo, Alexander Watts, David Fisman, Kamran Khan

## Abstract

The Coronavirus Disease 2019 (COVID-19) epidemic began in Wuhan, China in late 2019 and continues to spread globally, with exported cases confirmed in 28 countries at the time of writing. During the interval between February 19 and 23, 2020, Iran reported its first 43 cases with eight deaths. Three exported cases originating in Iran were identified, suggesting a underlying burden of disease in that country than is indicated by reported cases. A large epidemic in Iran could further fuel global dissemination of COVID-19. We sought to estimate COVID-19 outbreak size in Iran based on known exported case counts and air travel links between Iran and other countries, and to anticipate where infections originating in Iran may spread to next. We assessed interconnectivity between Iran and other countries using using International Air Transport Association (IATA) data. We used the methods of Fraser et al. to estimate the size of the underlying epidemic that would result in cases being observed in the United Arab Emirates (UAE), Lebanon, and Canada. Time at risk estimates were based on a presumed 6 week epidemic age, and length of stay data for visitors to Iran derived from the United Nations World Tourism Organization (UNWTO). We evaluated the relationship between the strength of travel links with Iran, and destination country rankings on the Infectious Disease Vulnerability Index (IDVI), a validated metric that estimates the capacity of a country to respond to an infectious disease outbreak. Scores range between 0-1, with higher scores reflecting greater capacity to manage infectious outbreaks. UAE, Lebanon, and Canada ranked 3rd, 21st, and 31st, respectively, for outbound air travel volume from Iran in February 2019. We estimated that 18,300 (95% confidence interval: 3770 to 53,470) COVID-19 cases would have had to occur in Iran, assuming an outbreak duration of 1.5 months in the country, in order to observe these three internationally exported cases reported at the time of writing. Results were robust under varying assumptions about undiagnosed case numbers in Syria, Azerbaijan and Iraq. Even if it were assumed that all cases were identified in all countries with certainty, the "best case" outbreak size was substantial (1820, 95% CI: 380-5320 cases), and far higher than reported case counts. Given the low volumes of air travel to countries with identified cases of COVID-19 with origin in Iran (such as Canada), it is likely that Iran is currently experiencing a COVID-19 epidemic of significant size for such exportations to be occurring. This is concerning, both for public health in Iran itself, and because of the high likelihood for outward dissemination of the epidemic to neighbouring countries with lower capacity to respond to infectious diseases epidemics.

## Background

The Coronavirus Disease 2019 (COVID-19) epidemic began in Wuhan, China in late 2019 and continues to spread globally (1), with exported cases confirmed in 28 countries at the time of writing (2). During the interval between February 19 and 23, 2020, Iran reported its first 43 cases with eight deaths. Three exported cases originating in Iran were identified, suggesting a underlying burden of disease in that country than is indicated by reported cases. A large epidemic in Iran could further fuel global dissemination of COVID-19.

## Objective

To quantify the COVID-19 outbreak size in Iran based on known exported case counts and air travel links between Iran and other countries, and to anticipate where infections originating in Iran may spread to next.

## Methods

We assessed interconnectivity between Iran and other countries using direct and total traveller volumes and final destination cities of travellers originating in Iran in February 2019, using data from the International Air Transport Association (IATA) (accounting for 90% of global air travel, with the other 10% modeled using market intelligence). As exported cases were identified in the United Arab Emirates (UAE), Lebanon, and Canada, we used the methods of Fraser et al (3) to estimate the size of the underlying epidemic in Iran necessary in order for these cases to be observed with a reasonable probability. To estimate the time at risk of COVID-19 exposure for travelers departing Iran, we obtained data from the United Nations World Tourism Organization (UNWTO) for the proportion of international travelers that are residents of Iran (4) and the average length of stay of tourists to Iran (5), and assumed that the Iranian outbreak began in early January 2020. We evaluated the relationship between the strength of travel links with Iran, and destination countries’ ranking on the Infectious Disease Vulnerability Index (IDVI), a validated metric that estimates the capacity of a country to respond to an infectious disease outbreak. Scores range between 0-1, with higher scores reflecting greater capacity to manage infectious outbreaks.

## Findings

We evaluated travellers from Iranian airports (Tehran, Rasht, and Arak) to international destinations in February 2019. While Qom has reported cases, the international airport is currently being built. Global cities receiving the greatest number of total travellers from Iran include Istanbul, Turkey (46,550), Najaf, Iraq (24,659), and Dubai, UAE (16,340) during this time. Among the top 10 traveller-receiving cities, four (Najaf, Baghdad, Damascus, and Baku) are located in countries with IDVI < 0.6, suggesting elevated vulnerability to infectious disease outbreaks as well as limited ability to detect cases (**Figure 1** and **Supplementary Table 1**).

**Figure 1.**
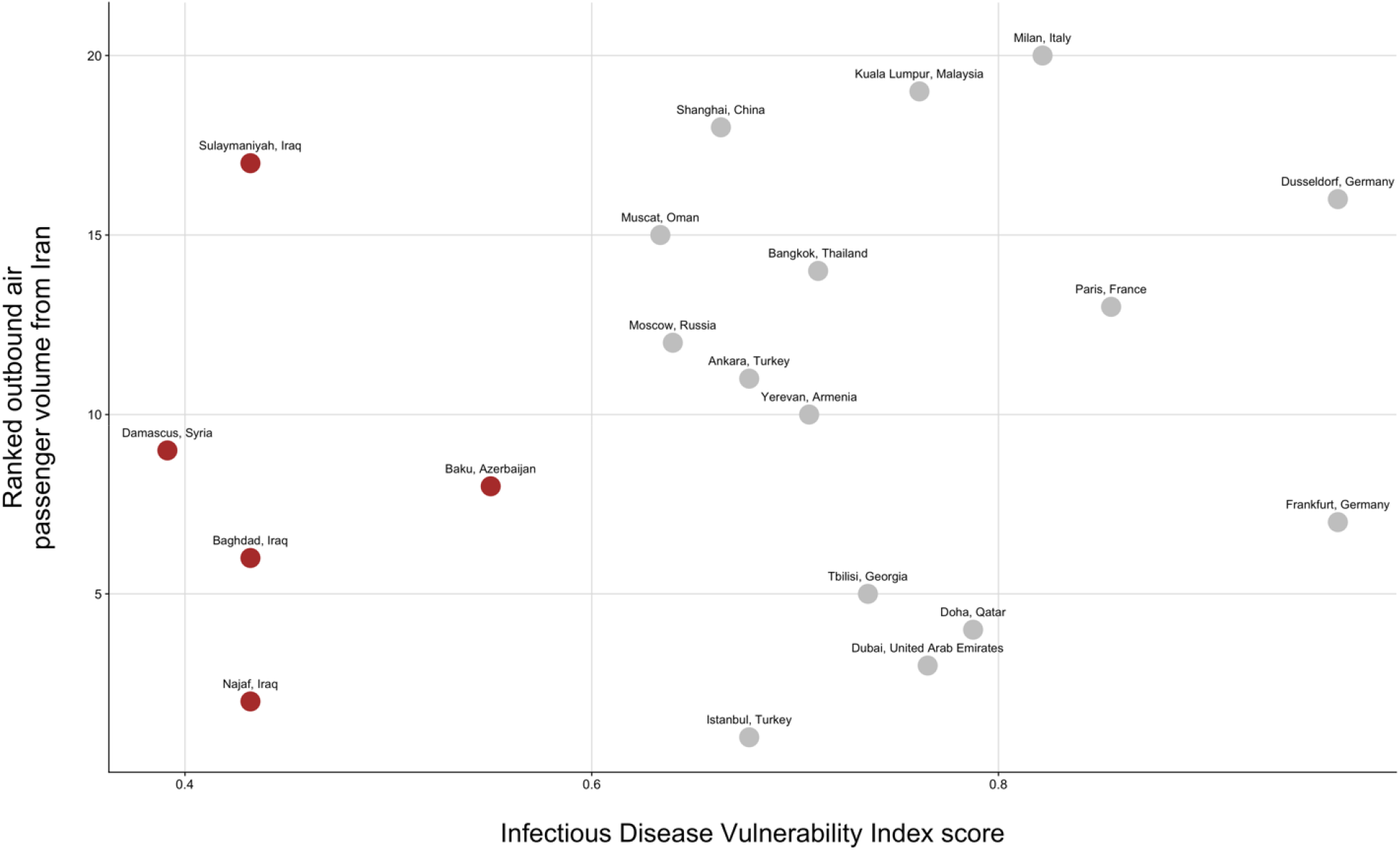
Top 20 international cities connected to Iran by commercial air travel and associated vulnerability to infectious disease outbreaks. Vulnerability is measured by the country-level using the Infectious Disease Vulnerability Index score, with a lower value indicating reduced capacity to respond to outbreaks. Countries with the lowest IDVI score are indicated in red. The top 20 cities accounted for 70% of international outbound traveller volumes from Iran in February 2019.

UAE, Lebanon, and Canada ranked 3^rd^, 21^st^, and 31^st^, respectively, for outbound air travel volume from Iran in February 2019. We estimated that 18,300 (95% confidence interval: 3770 – 53,470) COVID-19 cases would have had to occur in Iran, assuming an outbreak duration of 1.5 months in the country, in order to observe these three internationally exported cases reported at the time of writing.

Given the low rankings for Lebanon and Canada for outbound air travel, it is unexpected that cases should be identified in these countries, but not Iraq, Syria, or Azerbaijan (countries with higher travel volumes but low IDVI scores). Considering traveller volume alone, the odds of a single case being imported into Iraq rather than Canada or Lebanon would be 33.6:1 and 15.4:1 respectively; for Azerbaijan, the odds would be 3.8:1 and 1.7:1, respectively; while for Syria, the odds would be 3.7:1 and 1.7:1 respectively. As such, we performed exploratory analyses in which we assumed that an unidentified exported case of COVID-19 was present in Iraq, Syria, Azerbaijan, or all 3 countries, in addition to Lebanon, Canada and UAE and estimated the outbreak size in Iran that would produce these results (**Figure 2**). We also evaluated a scenario where we assumed perfect case detection in travelers from Iran, such that disease is truly absent in countries not reporting cases. Under this ‘best-case’ scenario, the estimated outbreak size in Iran was smaller but still substantial (1820, 95% CI: 380-5320 cases).

**Figure 2.**
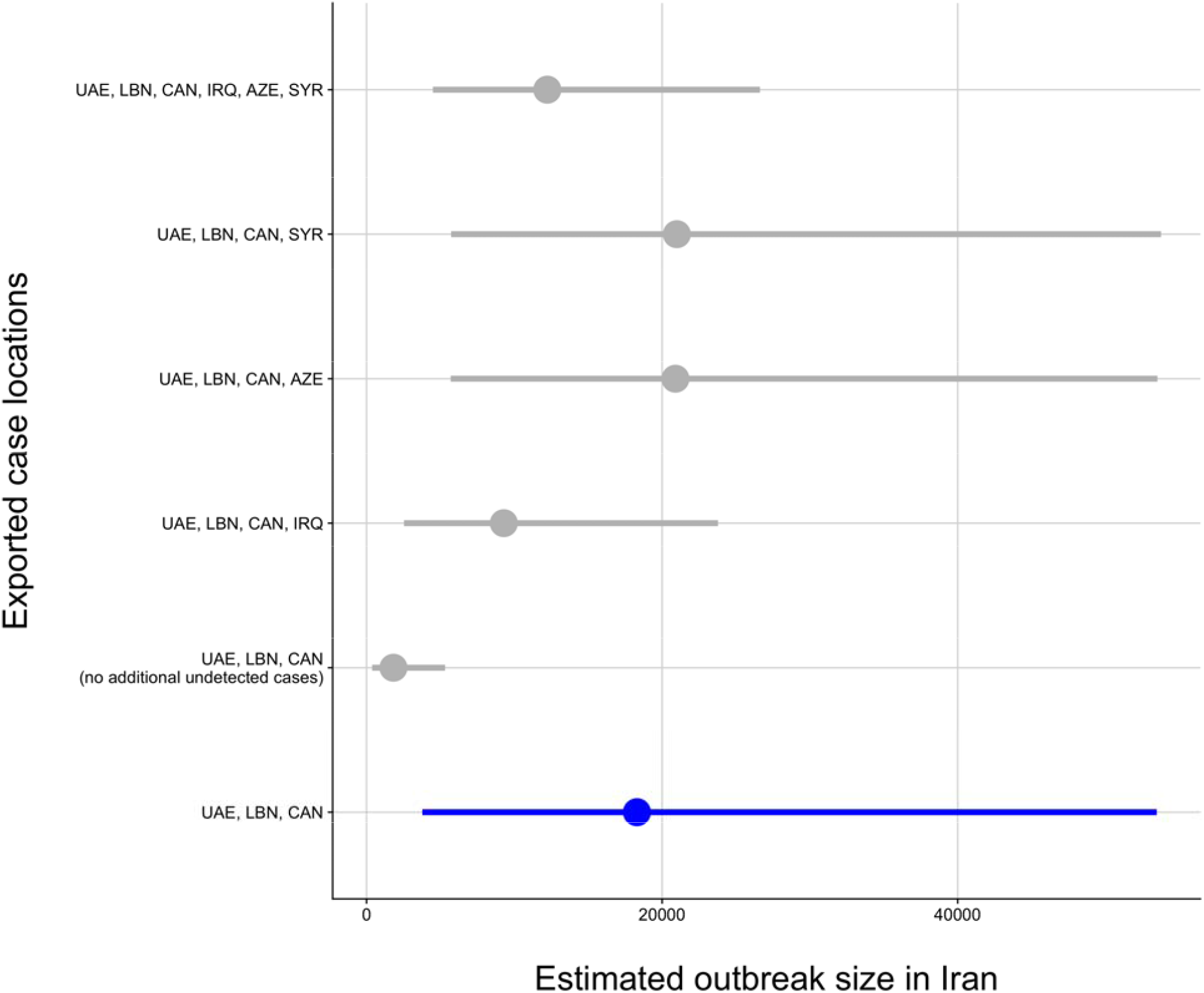
Estimated outbreak size in Iran required to observe exported cases internationally. The estimated cumulative number of COVID-19 cases in Iran required to observe 3 exported cases to UAE, LBN, and CAN is shown in blue. We also estimated the outbreak size required under alternate scenarios, including no additional exported cases to any other international destinations despite perfect case detection, and one additional exported case to IRQ, AZE, or SYR, (independently or to all 3 countries). Mean and 95% confidence intervals are presented. Abbreviations: UAE – United Arab Emirates; LBN – Lebanon; CAN – Canada; IRQ – Iraq; AZE – Azerbaijan; SYR – Syria.

## Discussion

Given the low volumes of air travel to countries with identified cases of COVID-19 with origin in Iran (such as Canada), it is likely that Iran is currently experiencing a COVID-19 epidemic of significant size for such exportations to be occurring. Our analysis would be modified by travel restrictions from Iran due to recent political situations, and by variations in the R0 value. Further, the lack of identified COVID-19 cases in countries with far closer travel ties to Iran suggests that cases in these countries are likely being missed, rather than being truly absent. This is concerning, both for public health in Iran itself, and because of the high likelihood for outward dissemination of the epidemic to neighbouring countries with lower capacity to respond to infectious diseases epidemics. Supporting capacity for public health initiatives in the region is urgently needed.

## Data Availability

Data used for this analysis are included in Supplementary Table 1 and are available via the URLs below.

https://www.nationsencyclopedia.com/WorldStats/UNCTAD-average-length-stay-visitors.html

---

**Supplementary Table 1.** Top 25 traveller destination cities from Tehran, Rasht, and Arak, Iran and corresponding Infectious Disease Vulnerability Index value for destination countries.

|  | **Destination city** | **Destination Country** | **Direct Travel Volume\*** | **Total Travel Volume\*** | **IDVI** |
| --- | --- | --- | --- | --- | --- |
| **1** | Istanbul | Turkey | 46403 | 46550 | 0.68 |
| **2** | Najaf | Iraq | 24612 | 24659 | 0.43 |
| **3** | Dubai | United Arab Emirates | 16340 | 16531 | 0.77 |
| **4** | Doha | Qatar | 5196 | 5223 | 0.79 |
| **5** | Tbilisi | Georgia | 4818 | 5067 | 0.74 |
| **6** | Baghdad | Iraq | 4305 | 4346 | 0.43 |
| **7** | Frankfurt | Germany | 3543 | 4240 | 0.97 |
| **8** | Baku | Azerbaijan | 3910 | 3910 | 0.55 |
| **9** | Damascus | Syria | 3799 | 3799 | 0.39 |
| **10** | Yerevan | Armenia | 3793 | 3793 | 0.71 |
| **11** | Ankara | Turkey | 2995 | 3611 | 0.68 |
| **12** | Moscow | Russia | 3102 | 3534 | 0.64 |
| **13** | Paris | France | 2967 | 3328 | 0.86 |
| **14** | Bangkok | Thailand | 2729 | 2982 | 0.71 |
| **15** | Muscat | Oman | 2535 | 2913 | 0.63 |
| **16** | Dusseldorf | Germany | 1806 | 2871 | 0.97 |
| **17** | Sulaymaniyah | Iraq | 2750 | 2841 | 0.43 |
| **18** | Shanghai | China | 2572 | 2812 | 0.66 |
| **19** | Kuala Lumpur | Malaysia | 2422 | 2683 | 0.76 |
| **20** | Milan | Italy | 2108 | 2503 | 0.82 |

\*Data from the International Air Travel Association (IATA), February, 2019.

